# FGF23 is a potential prognostic biomarker in uterine sarcoma

**DOI:** 10.1101/2023.08.20.23294198

**Authors:** Ling Yang, Ying Cai, Yunjia Wang, Yue Huang, Chi Zhang, Hu Ma, Jian-Guo Zhou

## Abstract

**Background:** Uterine sarcoma (US) is a highly malignant cancer with poor prognosis and high mortality in women. In this study, we evaluated the expression of human fibroblast growth factor 23 (FGF23) in different US subtypes and the relationship between survival and clinicopathological characteristics.

**Methods:** We conducted a comparative analysis of FGF23 gene expression in different pathological types of US. Utilizing a cohort from The Cancer Genome Atlas (TCGA) of 57 patients, a 50-patient microarray dataset (GSE119043) from the Gene Expression Omnibus (GEO) and a clinical cohort of 44 patients, we analyzed gene expression profiles and corresponding clinicopathological information. Immunohistochemistry (IHC) was used to examine the expression level of FGF23 in four US subtypes. Survival analysis was used to assess the relationship between FGF23 expression and prognosis in US patients.

**Results:** Compared with uterine normal smooth muscle (UNSM) and uterine leiomyoma (ULM), FGF23 expression was significantly upregulated in US and was differentially expressed in four US subtypes. Uterine carcinosarcoma (UCS) exhibited the highest expression of FGF23 among the subtypes. Survival analysis indicated that FGF23 expression was unrelated to overall survival and progression-free survival in US (P>0.05). Similar results were obtained from the validation cohorts. Univariate and multivariate analyses showed no significant correlation between FGF23 expression and the US prognosis. Tumor stage, CA125 and tumor recurrence were independent prognostic factors for survival of US patients.

**Conclusion:** FGF23 was highly expressed in uterine sarcoma and was promising as a novel potential biomarker for the diagnosis and prognosis of uterine sarcoma.

## Introduction

Uterine sarcoma (US) is one of the most aggressive and lethal gynecologic malignancies, comprising approximately 1% of all female reproductive tract malignancies and 3∼7% of all uterine cancers[1]. At present, the prognosis of US patients is extremely poor due to the advanced disease stage at diagnosis, which is caused by its nonspecific early symptoms that are similar to those of uterine myoma or adenomyosis. More than 50% of all US cases are diagnosed by chance[2]. Therefore, novel potential cost-effective biomarkers and powerful treatments are needed to improve diagnosis and prognosis in cases of US. Nevertheless, the discovery of new biomarkers and specific targeted therapies is still challenging owing to the low incidence of the disease[3].

Human fibroblast growth factor 23 (FGF23) is a member of the fibroblast growth factor (FGF) family[4]. It is secreted by bone cells and osteoblasts and is responsible for regulating phosphorus levels in a hormone-like manner. Studies have revealed that FGF23 is closely related to gynecological tumors. Serum or plasma FGF23 concentrations were significantly elevated in women with advanced-stage ovarian cancer compared to women with early-stage ovarian cancer, benign disease or healthy women. Additionally, a positive correlation was observed between FGF23 and the stage of the disease[5]. Moreover, a distinct single-nucleotide polymorphism (SNP) of FGF23 exhibited a significant correlation with treatment response following platinum-based chemotherapy and surgery as well as the overall survival of patients with ovarian cancer[6]. Nonetheless, the expression of FGF23 in US remains unclear. In this study, to clarify the role of FGF23 in US, we explored the relationship between FGF23 expression and the prognosis of US.

## Materials and Methods

### Data resource

This study included three cohorts in total. The gene expression profiles and corresponding clinicopathological information were downloaded from two open databases, including a cohort from The Cancer Genome Atlas (TCGA) data portal (http://cancergenome.nih.gov/) and a microarray dataset (GSE119043) from the Gene Expression Omnibus (GEO) (https://www.ncbi.nlm.nih.gov/geo/), which contained data from 57 UCS patients and 50 UUS patients, respectively. The above data are publicly available. A total of 44 diagnosed patients and clinicopathological information were collected from the Department of Pathology of Suining Central Hospital from May 2014 to January 2022. This study received approval from the Review Board of Suining Central Hospital and the Ethics Committee of Zunyi Medical University.

The primary endpoint was overall survival (OS), defined as days from initial diagnosis to death or last follow-up. Progression-free survival (PFS) was defined as days from initial diagnosis to disease progression or death, whichever occurred first (or the date of last follow-up if progression or death had not yet occurred). Staging was assessed according to the International Federation of Gynecology and Obstetrics (FIGO) stage system[7].

### Pathological inclusion criteria

According to the the National Comprehensive Cancer Network (NCCN) guidelines[8], US pathological types are most commonly include leiomyosarcoma (LMS), endometrial stromal sarcoma (ESS), undifferentiated uterine sarcoma (UUS) and adenosarcoma (AS). Although uterine carcinosarcoma (UCS) is classified as a dedifferentiated or metaplastic form of endometrial carcinoma, due to its more aggressive behavior compared with the usual type of endometrial carcinoma, carcinosarcoma is still included in most retrospective studies of US as well as in the separate section of “mixed epithelial and mesenchymal tumors” of the 2014 WHO classification[9]. Therefore, we included patients definitely pathologically diagnosed with LMS, ESS, AS, UUS and UCS in this study. Patients with biopsy-proven uterine sarcoma per the 2020 WHO classification of uterine sarcoma were included[10]. Patients without a definitive histologic diagnosis, patients with metastatic sarcoma from other gynecological sides were excluded from the study. Patients with incomplete information regarding pathological diagnosis, clinical findings, and follow-up studies for analysis, as they were lost to follow-up after initial diagnosis, were also excluded.

### Differentially expressed analysis

To compare the differential expression of FGF23 genes between tumor tissues and normal tissues in US patients, FGF23 gene expression was analyzed from the TCGA cohort according to the parameters |log_2_-fold change| >1 and P value < 0.05.

### Immunohistochemistry (IHC)

In this study, we thoroughly reviewed the medical records of 44 patients with pathologically confirmed US, including 16 samples of LMS, 14 of ESS, 13 of UCS, and 1 of AS, in the Department of Pathology of Suining Central Hospital. Smooth muscle tissue of the uterine wall and leiomyoma tissue were also obtained if present.

Immunohistochemistry was performed using a mouse anti-human FGF23 monoclonal antibody (NBP3-07378; Novus Biologicals, Colorado, USA). Tissue specimens were fixed with 10% formalin and preserved in paraffin. The paraffin-embedded blocks were sectioned continuously to a thickness of 4 µm and fixed on slides for immunohistochemical staining. Paraffin-embedded sections were dewaxed and then repaired using ethylenediaminetetraacetic acid (EDTA) antigen retrieval buffers. The primary antibody was used at a 1:100 dilution with an incubation of 2 hours at room temperature. Secondary antibody and detection were performed using an automated immunostainer (VENTAN Bench Mark XT, Roche, Swiss), followed by counterstaining with hematoxylin. Normal human kidney tissue was used as a positive control. The higher the expression content of FGF23 was, the greater was the distribution density and the stronger the positive result would be.

FGF23 expression was evaluated using the H-score, which is defined as the product of the staining intensity and the percentage of positively stained cells. The scoring system for staining intensity was as follows: 0 points for no staining, 1 point for light yellow, 2 points for brown□yellow, and 3 points for brown. The percentage of positive cells was also considered, and the scores were as follows: 0 points, positive cells 5% or less; 1 point, positive cells between 6% and 25%; 2 points, positive cells between 26% and 50%; 3 points, positive cells between 51%-75%; and 4 points, positive cells greater than 75%. The resulting total scores were categorized as negative (0 points), weakly positive (1-4 points), moderately positive (5-8 points), and strongly positive (9-12 points). The scoring process was performed independently by two gynecologic pathologists using optical microscopy. The decision was made after discussion within the group if inconsistent.

### Statistical analysis

The statistical analysis was conducted using IBM® SPSS® Statistics software version 24 (IBM, Armonk, NY), R (version 4.2.2) and GraphPad Prism 9.5.1. The distributions of categorical variables and continuous variables between cases and controls were performed using Pearson ^χ2^ tests and Student’s t tests, respectively. Categorical data are presented as counts and percentages and were analyzed using the Chi-square test and Fisher’s exact test. The skewed distributions were described with medians and interquartile ranges (IQR). The expression levels of FGF23 in patients were determined using the *ggplot2* package.

Kaplan-Meier analysis was used to evaluate the relationship between overall survival and FGF23 expression level and pathological subtypes in the US. Univariate and multivariate Cox proportional hazard regression analyses were used to investigate FGF23 expression as an independent prognostic factor of US. P<0.05 was considered statistically significant.

## Results

### Differentially expressed analysis

A cohort of 57 UCS patients and corresponding necessary clinical data were downloaded from the TCGA database. We analyzed FGF23 expression levels between tumor tissues and normal tissues in 57 UCS samples and found that there was no significant difference in FGF23 gene expression between tumor and normal tissues (P > 0.005; Supplementary Fig. S1).

### Baseline characteristics

According to the guidelines of the FIGO, the clinical data collected from the patients’ medical records were analyzed, including age at diagnosis, histologic types, clinical and pathologic staging, and adjuvant chemotherapy (CT) or radiation treatment (RT). These baseline characteristics related to two cohorts, which were from the TCGA database, including 57 UCS patients, and a clinical cohort including 44 US cases from the Department of Pathology of Suining Central Hospital. Supplementary Table S1 shows the baseline characteristics of 57 patients in the TCGA cohort. All patients involved in the study were female and over age 50. Most of them (n=56, 98.25%) were postmenopausal. Regarding race, most of the patients (n=44, 77.19%) were White. Peritoneal wash included positive (n=12, 21.05%), negative (n=25, 43.86%) and unknown (n=20, 35.09%). The FIGO stage included 21 (36.84%) I, 6 (10.53%) II, 20 (35.09%) III and 10 (17.54%) IV. Surgical approaches included minimally invasive (n=30, 52.63%), open (n=20, 35.09%) and unknown (n=7, 12.28%). Person neoplasm cancer status included 9 (15.79%) with tumors, 11 (19.30%) tumor free and 37 (64.91%), unknown. In terms of adjuvant treatment, 11 patients received radiotherapy (RT, 19.30%), 10 patients (17.54%) never used tamoxifen, and 47 patients unknown (82.46%).

From an initial cohort of 56 patients, 44 cases were identified with a definite pathological subtype of uterine sarcoma. The baseline characteristics of 44 US patients from the clinical(Suining) cohort are presented in Table 1.

**Table 1.** Baseline characteristics of 44 US patients from the Suining cohort.

| Characteristics | All Patients<br>(n=44) | AS<br>(n=1) | UCS<br>(n=13) | ESS<br>(n=14) | LMS<br>(n=16) | P |
| --- | --- | --- | --- | --- | --- | --- |
| **Age at diagnosis(years), n(%)** |  |  |  |  |  | 0.539 |
| ≤50 | 21 (47.73) | 1 (4.76) | 5 (23.81) | 6 (28.57) | 9 (42.86) |  |
| > 50 | 23 (52.27) | 0 (0.0) | 8 (34.78) | 8 (34.78) | 7 (30.43) |  |
| **Menopausal status, n(%)** |  |  |  |  |  | 0.202 |
| Postmenopausal | 20 (45.45) | 0 (0.0) | 9 (45.0) | 5 (25.0) | 6 (30.0) |  |
| Premenopausal | 24 (54.55) | 1 (4.17) | 4 (16.67) | 9 (37.5) | 10 (41.67) |  |
| **Symptoms, n(%)** |  |  |  |  |  | 0.010 |
| Colporrhagia | 20 (45.45) | 0 (0.0) | 11 (55.0) | 6 (30.0) | 3 (15.0) |  |
| Stomachache/bloating | 7 (15.91) | 0 (0.0) | 0 (0.0) | 2 (28.57) | 5 (71.43) |  |
| Abdominal mass | 11 (25.0) | 0 (0.0) | 1 (9.09) | 4 (36.36) | 6 (54.55) |  |
| Asymptomatic | 6 (13.64) | 1 (16.67) | 1 (16.67) | 2 (33.33) | 2 (33.33) |  |
| **Tumor size(cm), n(%)** |  |  |  |  |  | 0.062 |
| ≤5 | 16 (36.36) | 1 (6.25) | 6 (37.5) | 7 (43.75) | 2 (12.5) |  |
| > 5 | 28 (63.64) | 0 (0.0) | 7 (25.0) | 7 (25.0) | 14 (50.0) |  |
| **CA125(U/ml), n(%)** |  |  |  |  |  | 0.679 |
| < 35 | 18 (40.91) | 1 (5.56) | 4 (22.22) | 6 (33.33) | 7 (38.89) |  |
| > 35 | 14 (31.82) | 0 (0.0) | 6 (42.86) | 3 (21.43) | 5 (35.71) |  |
| Unknown | 12 (27.27) | 0 (0.0) | 3 (25.0) | 5 (41.67) | 4 (33.33) |  |
| **FIGO stage, n(%)** |  |  |  |  |  | 0.352 |
| □ | 27 (61.36) | 1 (3.7) | 5 (18.52) | 10 (37.04) | 11 (40.74) |  |
| □ | 6 (13.64) | 0 (0.0) | 3 (50.0) | 2 (33.33) | 1 (16.67) |  |
| □ | 5 (11.36) | 0 (0.0) | 3 (60.0) | 2 (40.0) | 0 (0.0) |  |
| □ | 6 (13.64) | 0 (0.0) | 2 (40.0) | 0 (0.0) | 4 (60.0) |  |
| **Surgical approach, n(%)** |  |  |  |  |  | 0.072 |
| TH | 1 (2.27) | 0 (0.0) | 1 (100.0) | 0 (0.0) | 0 (0.0) |  |
| TH+BSO | 24 (54.55) | 0 (0.0) | 8 (33.33) | 8 (33.33) | 8 (33.33) |  |
| TH+BSO+PLA | 11 (25.0) | 1 (9.09) | 4 (36.36) | 5 (45.45) | 1 (9.09) |  |
| TH+BSO+PLA+omentectomy | 5 (11.36) | 0 (0.0) | 0 (0.0) | 1 (20.0) | 4 (80.0) |  |
| TH+BSO+PLA+omentectomy+appendecto | 3 (6.82) | 0 (0.0) | 0 (0.0) | 0 (0.0) | 3 (100.0) |  |
| my |  |  |  |  |  |  |
| **Lymphadenectomy, n(%)** |  |  |  |  |  | 0.511 |
| Yes | 19 (43.18) | 1 (5.26) | 4 (21.05) | 6 (31.58) | 8 (42.11) |  |
| No | 25 (56.82) | 0 (0.0) | 9 (36.0) | 8 (32.0) | 8 (32.0) |  |
| **Lymphatic metastasis, n(%)** |  |  |  |  |  | < 0.001 |
| Yes | 3 (6.82) | 0 (0.0) | 3 (100) | 0 (0.0) | 0 (0.0) |  |
| No | 16 (36.36) | 1 (6.25) | 1 (6.25) | 6 (37.5) | 8 (50.0) |  |
| Unknown | 25 (56.82) | 0 (0.0) | 9 (36.0) | 8 (32.0) | 8 (32.0) |  |
| **Chemotherapy, n(%)** |  |  |  |  |  | 0.326 |
| Yes | 22 (50.0) | 0 (0.0) | 9 (40.91) | 6 (27.27) | 7 (31.82) |  |
| No | 22 (50.0) | 1 (4.55) | 4 (18.18) | 8 (36.36) | 9 (40.91) |  |
| **Radiotherapy, n(%)** |  |  |  |  |  |  |
| Yes | 11 (25.0) | 0 (0.0) | 5 (45.45) | 2 (18.18) | 4 (36.36) | 0.485 |
| No | 33 (75.0) | 1 (3.03) | 8 (24.24) | 12 (36.36) | 12 (36.36) |  |

TH: Total hysterectomy; TH+BSO: Total hysterectomy+bilateral salpingo-oophorectomy; PLA: Pelvic lymphadenectomy

The mean age of all participants was 51.5 ±10 years (range, 35 to 73 years). Regarding menopausal status, 20 (45.45%) patients were postmenopausal, and 24 (54.55%) patients were premenopausal. The symptoms included colporrhagia (n=20, 45.45%), stomachache/bloating (n=7, 15.91%), abdominal mass (n=11, 25.00%) and asymptomatic (n=6, 13.64%). In terms of CA125 levels, 14 (31.82%) patients had CA125 levels >35 U/ml, 18 (40.91%) patients had CA125 levels <35 U/ml, and 12 (27.27%) patients were unknown. The FIGO stage included 27 (61.36%) I, 6 (13.64%) II, 5 (11.36%) III and 6 (13.64%) IV. Surgical approaches included total hysterectomy (TH, n=1, 2.27%), TH+bilateral salpingo-oophorectomy (TH+BSO, n=24, 54.55%), TH+BSO+pelvic lymphadenectomy (TH+BSO+PLA, n=11, 25.00%), TH+BSO+PLA+omentectomy (n=5, 11.36%) and TH+BSO+PLA+omentectomy+appendectomy (n=3, 6.82%). A total of 43.18% (n=19) of all patients underwent lymphadenectomy, and 6.82% (n=3) experienced lymphatic metastasis. In addition, for adjuvant treatment, half of the patients (n=22, 50.0%) received chemotherapy (CT) with doxorubicin and ifosfamide as the primary protocol, while the rest did not. Eleven patients (25.0%) underwent radiotherapy (RT), while 33 patients (75.0%) did not. Forty-four patients were categorized according to pathological type into groups of adenosarcoma (AS, n=1, 2.27%), uterine carcinosarcoma (UCS, n=13, 29.55%), endometrial stromal sarcoma (ESS, n=14, 31.82%) (including 12 low-grade ESS (LG-ESS) and 2 high-grade ESS (HG-ESS)) and leiomyosarcoma (LMS, n=16, 36.36%).

### IHC Evaluation of FGF23

Immunohistochemistry was conducted to examine the expression of FGF23 in 44 full-face tissue sections obtained from US patients, including 32 paired samples of normal uterine smooth muscle (UNSM) and 6 paired samples of uterine leiomyoma (ULM). Positive expression of FGF23 protein was detected in the cell membrane and cytoplasm, appearing as light yellow, brownish yellow or dark brown particles. Notably, only one endometrial stromal sarcoma (ESS) sample displayed sporadic and subdued heterogeneous staining in 26-50% of cells, which was classified as weakly positive. Additionally, all specimens from US patients showed varying levels of positivity, ranging from moderate to strong.

As shown in Fig. 1, among 6 paired samples, the expression level of FGF23 exhibited a significant increase in the US group compared to both the UNSM and ULM groups (P=0.005 and P=0.006, respectively) and in the ULM group compared to the UNSM group (P=0.02). Furthermore, among 32 paired samples, similar results were obtained both in the US group compared to the UNSM group (P<0.001).

**Fig. 1.**
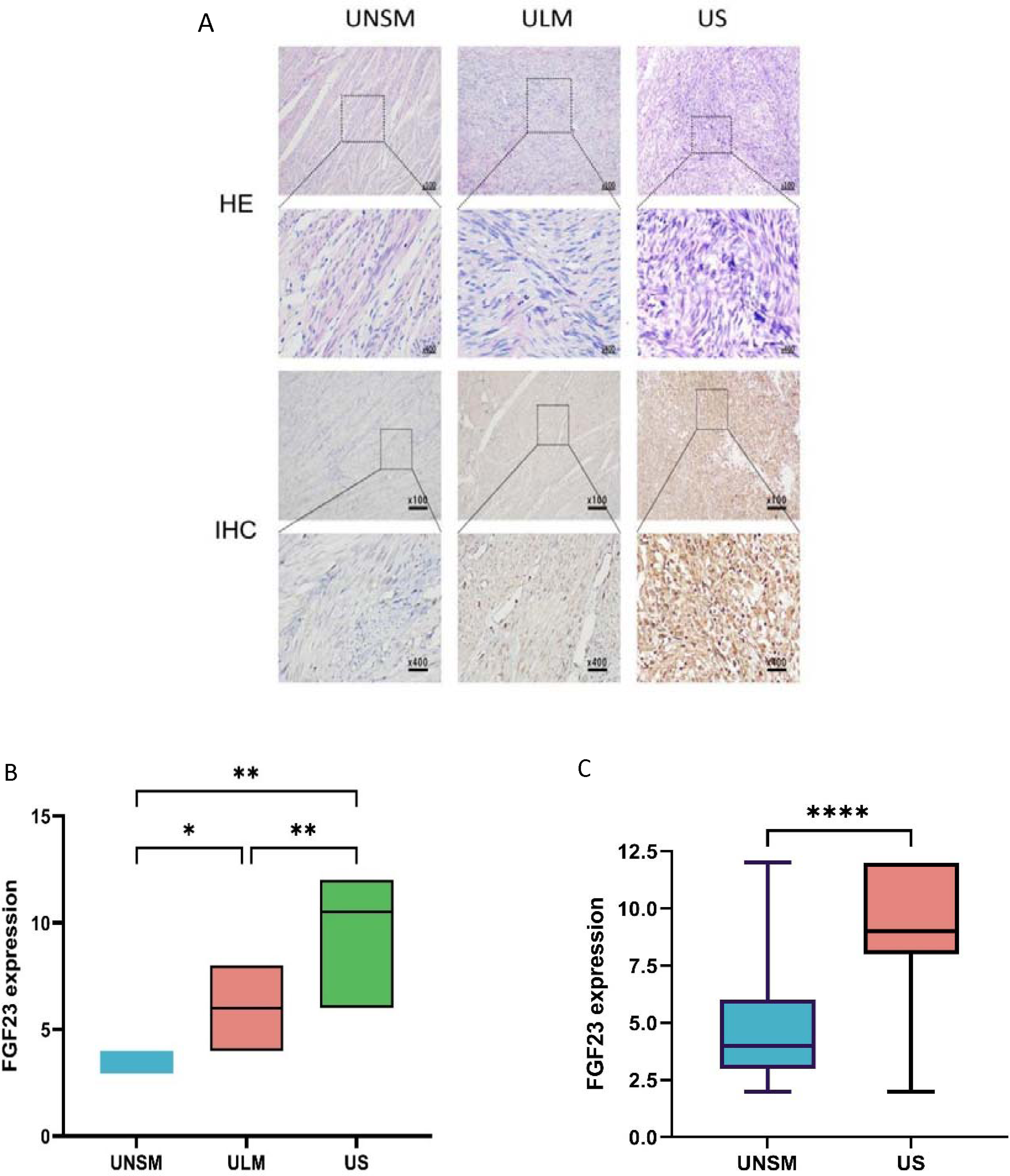
(A) IHC indicated the expression levels of FGF23 in the UNSM, ULM and US groups. (B) The box plot shows FGF23 expression in US, ULM, and UNSM among 6 paired samples. (C) The box plot shows FGF23 expression in US and UNSM among 32 paired samples.

As shown in Fig. 2, immunohistochemistry and statistical analysis revealed a significant difference (P=0.01) in FGF23 expression levels among AS, ESS, LMS and UCS. The expression of FGF23 in AS showed moderate positivity and was significantly lower than that in ESS, LMS and UCS, although there was only a single patient. Furthermore, the expression of FGF23 in ESS was substantially lower than that observed in LMS and UCS. Obviously, the findings indicated that UCS patients exhibited the highest levels of FGF23 expression when compared with other types (Fig. 2B).

**Fig. 2.**
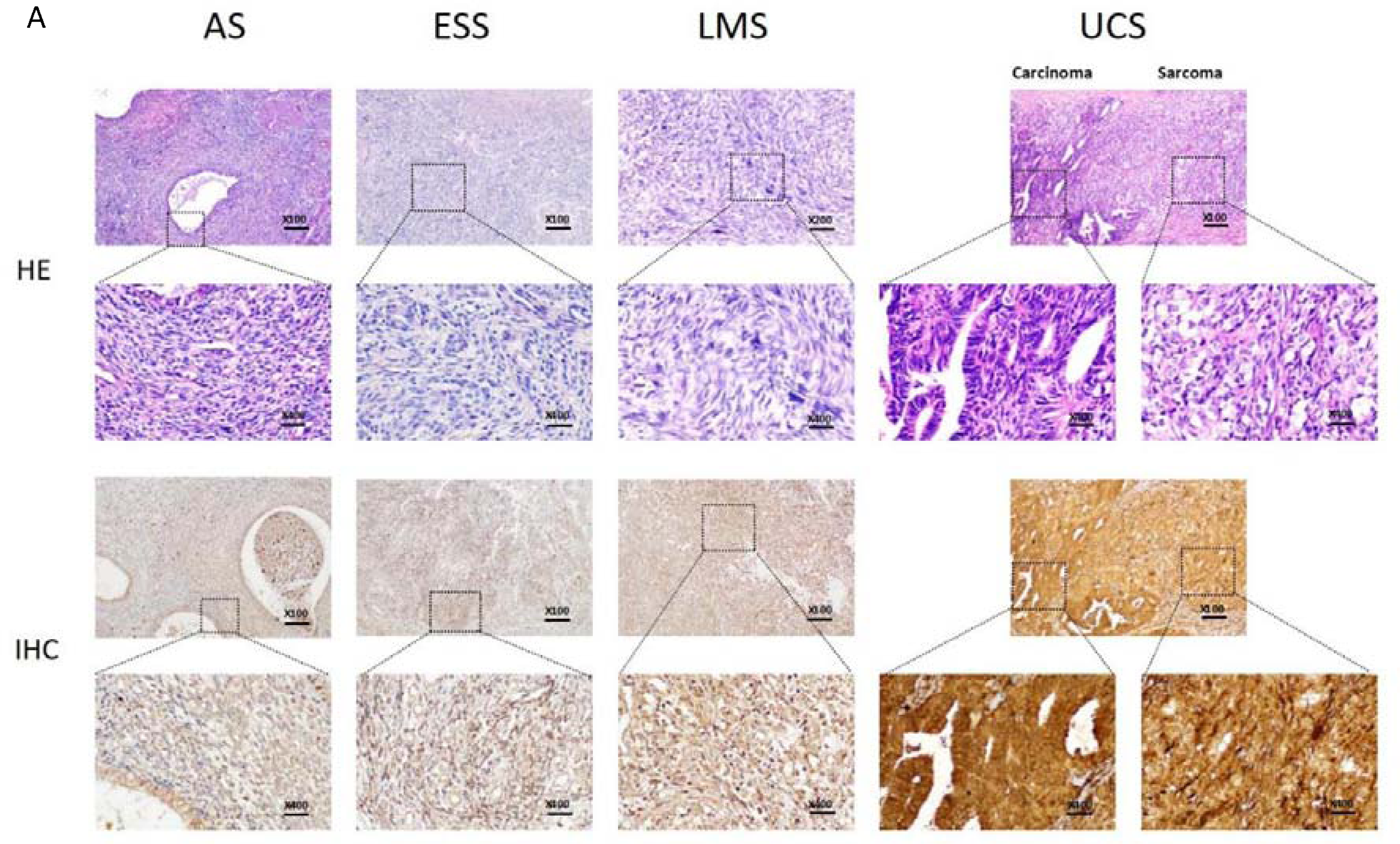
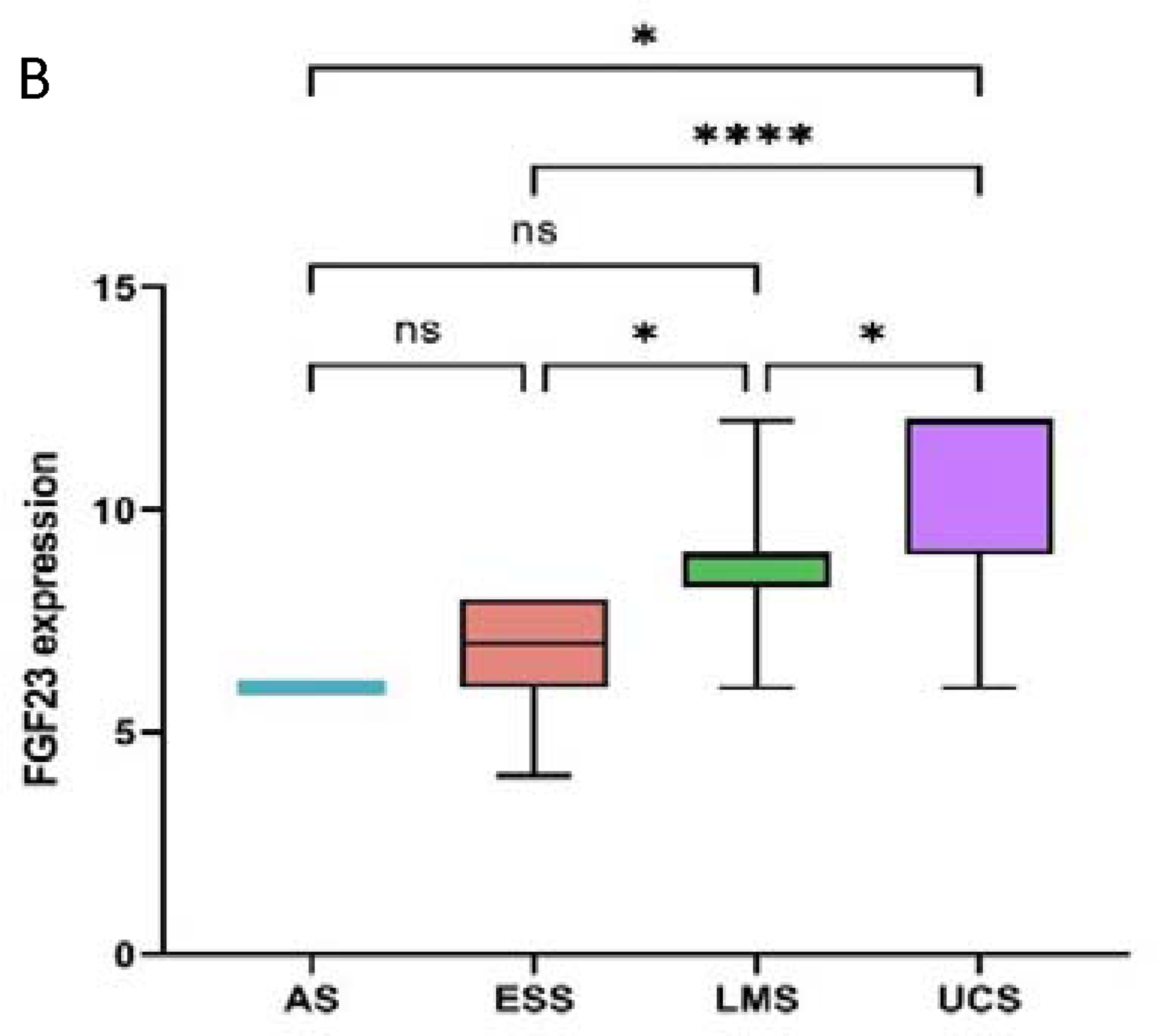
(A) IHC assay indicated FGF23 expression in different uterine sarcoma subtypes. (B) The box plot shows the differential expression of FGF23 in AS, ESS, LMS and UCS.

### Survival analysis

Based on the mean value of FGF23 expression, the patients were categorized into high-FGF23 and low-FGF23 expression groups, and we performed Kaplan-Meier analysis in three cohorts from TCGA, GEO and clinical cohorts. Kaplan-Meier analysis indicated that 41 UCS patients from the TCGA database with high FGF23 expression levels were unrelated to poor OS (P=0.22, HR: 2.01, 95% CI: 0.66-6.14; Fig. 3A), and the same results were obtained in 50 UUS patients from the GSE119043 microarray (P=0.065, HR: 1.87, 95% CI: 0.96-3.64; Fig. 3B). Then, the values of OS and PFS in the clinical cohort including 44 patients suggested similar results as shown in Fig. 3C (P=0.38, HR: 1.53, 95% CI: 0.59-3.99 and P=0.66, HR: 1.26, 95% CI: 0.45-3.51, respectively). The median OS time of patients with low FGF23 expression was not reached (NR), indicating a longer but not statistically significant survival compared to the median OS (48.7 months) in the high expression group. The median PFS times of patients with high or low FGF23 expression were not reached.

**Fig. 3.**
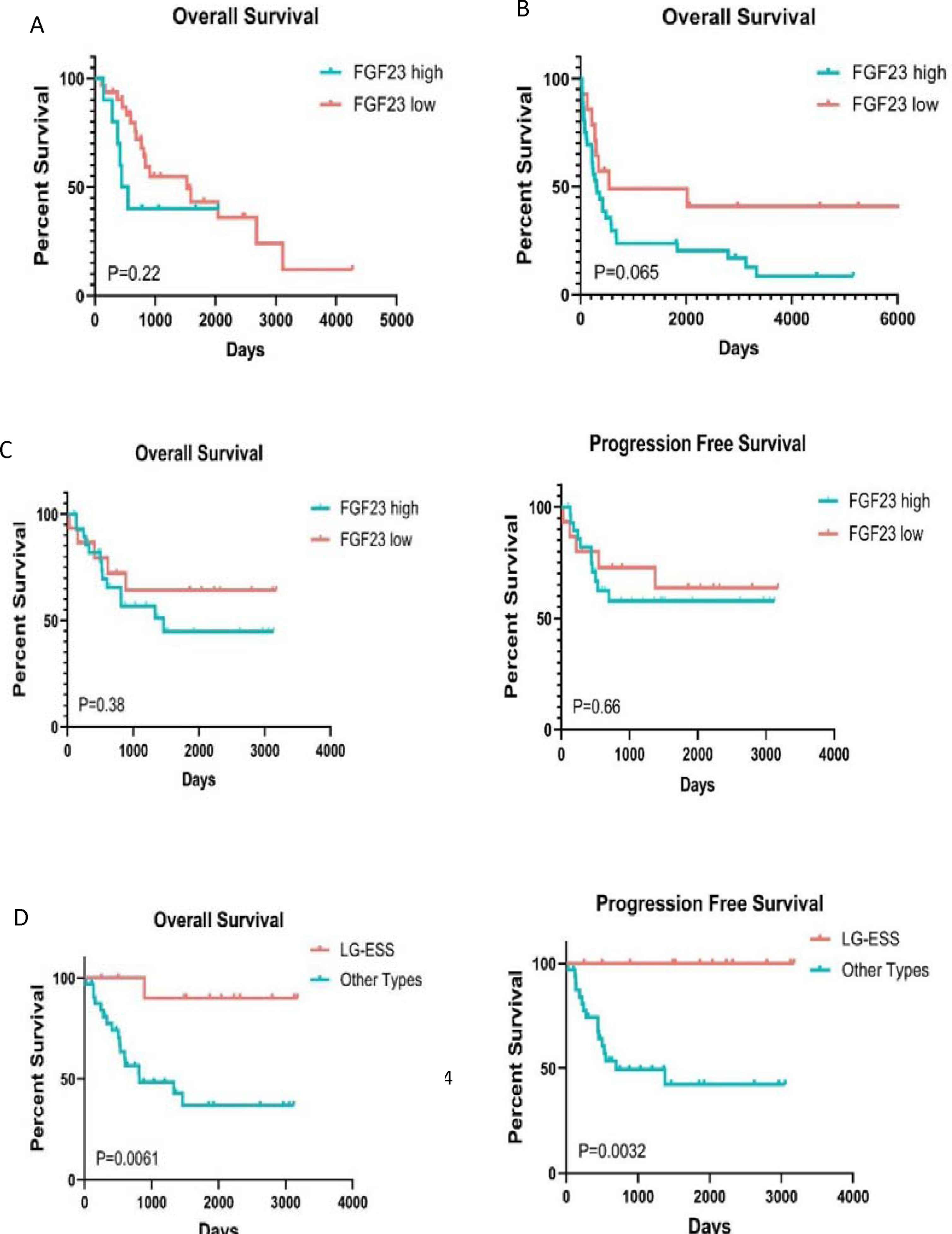
(A) K-M analysis suggested overall survival of FGF23 high or low expression in 41 UCS patients from the TCGA cohort. (B) K-M analysis revealed the overall survival by FGF23 high or low expression in 50 UUS patients from the GEO database. (C) K-M analysis indicated no significant correlation between FGF23 expression levels and OS or PFS of US patients from the clinical cohort. (D) K-M analysis showed the differential OS and PFS between LG-ESS and other types of US patients from the clinical cohort.

However, we analyzed the various pathological subtypes of 44 US patients from the clinical cohort and found that low-grade endometrial stromal sarcoma (LG-ESS) showed significantly better survival than other subtypes in terms of OS and PFS (P=0.006, HR: 0.26, 95% CI: 0.10-0.68 and P=0.003, HR: 0.21, 95% CI: 0.07-0.59, respectively; Figure 3D).

### Univariate and multivariate Cox regression analysis

Univariate and multivariate Cox proportional hazard regression analyses were carried out with UCS patients from the TCGA database. The univariate analysis indicated that tumor stage (HR=1.39; P=0.029; 95% CI: 1.03-1.87), neoplasm cancer status (HR=0.16; P=0.031; 95% CI: 0.03-0.85), peritoneal wash (HR=0.19; P=0.003; 95% CI: 0.07-0.57) and adjuvant radiotherapy (HR=0.46; P=0.006; 95% CI: 0.26-0.79) were significantly associated with OS in UCS patients. Multivariate analysis showed similar results (P<0.05). However, we found no significant correlation between FGF23 expression and OS (HR=1.06; P=0.532; 95% CI: 0.47-2.41). Furthermore, other clinical parameters, such as age, menopausal status, race, BMI and surgical approaches, were also unrelated to OS (P>0.05). The findings are presented in Supplementary Table S2.

Furthermore, we conducted univariate and multivariate Cox proportional hazard regression analyses of OS and PFS in US patients from the clinical cohort. As shown in Supplementary Table S3 and S4, the results revealed that tumor stage (HR=3.62; P=0.007; 95% CI: 1.42-9.22), CA125 (HR=3.23; P=0.001; 95% CI: 1.58-6.62), tumor type (HR=5.71; P=0.021; 95% CI: 1.30-25.13) and tumor recurrence (HR=8.47; P<0.001; 95% CI: 3.08-23.27) were significantly associated with OS in US patients, and also with PFS. Multivariate analysis showed that only CA125 and tumor recurrence acted as prognostic factors for OS, and only tumor recurrence acted as a prognostic factor for PFS (P<0.05). Nevertheless, the findings indicated that there was no correlation between FGF23 expression and OS (HR=1.58, P=0.383, 95% CI: 0.59-3.99) or PFS (HR=1.07, P=0.59, 95% CI: 0.88-1.29) in US patients, which was consistent with prior results from the TCGA cohort. Additionally, other clinical parameters, such as age, menopausal status, tumor size, lymphadenectomy, lymphatic metastasis, adjuvant chemotherapy and adjuvant radiotherapy, were also unrelated to OS and PFS (P>0.05).

## Discussion

US is one of the most aggressive soft tissue sarcomas (STSs) in adult women, with an increasing incidence and high mortality rate of up to 30% due to its highly aggressive nature[11]. Typically, this tumor is diagnosed in older women among whom the median age ranges from 62 to 67 years. Nonetheless, our study revealed a younger patient population, with a median age of 51.5 years and with a higher proportion of early-stage disease, which was observed in 75% (n=33) of patients versus 58-66% in the published literature[12].

Our results revealed that tumor stage, CA125, tumor type, tumor recurrence, neoplasm cancer status and peritoneal wash were significantly associated with OS in US patients. Other clinical parameters, such as age, menopausal status, race, BMI, surgical approaches, tumor size, lymphadenectomy, lymphatic metastasis and adjuvant radiotherapy, were unrelated to OS. Several previous researchers have confirmed that radiation therapy in patients with US can improve survival[13–15], we get the same result in TCGA cohort. However, the findings indicated that adjuvant radiotherapy did not improve the survival of US patients in clinical cohort compared with previous studies[16,17]. The role of radiation therapy in patients with US still needs further investigation [18]. In addition, our research showed the opposite outcome compared with a prior study, which found that poor survival was associated with old age and Black race[13]. Previous research confirmed that histological type and tumor size were independent prognostic factors for both OS and PFS, and tumor stage was significantly associated with PFS[19], our study exhibited similar results except for tumor size. However, there are no studies on the association between FGF23 expression and the prognosis of US at present.

In this study, we investigated the relationship between FGF23 expression and the clinical characteristics and survival of US patients. Consistently, tumor stage, neoplasm cancer status, peritoneal wash, CA125 and tumor recurrence were independent prognostic factors in US survival. LMS was the most prevalent histology in previous reports and was also observed in our investigation. At the time of analysis, 18 patients had died, and the three-year OS rate was 45.45% (n=20). Furthermore, our findings demonstrated that LG-ESS had longer PFS and OS than other pathological subtypes in the US, which is consistent with other published literature[13,20]. The principal surgical intervention, performed in 97.73% (n=43) of patients, was hysterectomy with bilateral salpingo-oophorectomy.

The rarity and heterogeneity of US presents tremendous challenges in identifying appropriate biomarkers and treatment modalities, resulting in limited progress in survival rates in recent decades[3]. Carcinogenesis is a multifaceted process involving numerous genes and factors. Among these, FGF23 and phosphate have been confirmed as cancer-related factors, with FGF23 being particularly relevant in bone-related cancers such as multiple myeloma or those with bone metastasis. Certain indications showed that oncogenic osteomalacia and elevated FGF23 levels may be relevant to breast cancer[21]. Breast cancer cells exhibit elevated FGF23 mRNA expression, and FGF secreted by malignant cells contributes to the formation of metastatic lesions[22]. In endometrial cancer, the median FGF23 concentrations in serum did not exhibit any notable distinctions when compared to benign endometrial changes[23]. Conversely, advanced-stage epithelial ovarian cancer (EOC) is characterized by an increase in FGF23 plasma concentration[5], and a specific FGF23 SNP has been related to a more favorable prognosis in this particular tumor type[6].

In our study, we analyzed the expression of FGF23 in US and its correlation with the prognosis of patients for the first time. We evaluated the expression of FGF23 in specimens of US patients using immunohistochemical assays. We found that FGF23 was expressed at low levels in UNSM and ULM but was highly expressed in US, while survival analysis in the TCGA, GEO and clinical cohorts showed that there was no significant correlation between the expression level of FGF23 and the prognosis of US patients. Moreover, by univariate and multivariate Cox regression analyses, we could not determine the statistical significance of FGF23 expression in OS. Considering that there were only 57 patients in the TCGA cohort, 50 patients in the GSE119043 microarray and 44 patients in the clinical cohort, the sample size was small and may not have afforded sufficient statistical efficacy; the median PFS and OS were not reached in the low FGF23 expression group at the end of the study, despite a median follow-up of 62.2 (range 1-105.9) months; therefore, the conclusions may be biased. Further study should be performed to verify the prognostic predictive role of FGF23 in uterine sarcoma.

Furthermore, in the clinical US subgroups, we found that LG-ESS showed significantly better OS and PFS than the other subtypes. This result was consistent with previous studies[24]. However, we did not analyze the difference between HG-ESS and LG-ESS because there were only two HG-ESS patients, which may have led to sampling bias. We did not measure the levels of FGF23 mRNA expression in the clinical cohort due to sample limitations, presenting another limitation in our study.

## Conclusion

FGF23 is significantly upregulated in uterine sarcoma and serves as a potentially novel diagnostic and prognostic biomarker.

## Ethics approval and consent to participate

This study was approved by the Review Board of Suining Central Hospital (No. LLSLH20220051) and the Ethics Committee of Zunyi Medical University (No. 2020-1-013) according to the guidelines of the Declaration of Helsinki. Informed consent was obtained from each participant.

## Authors’ contributions

Ling Yang: conceptualization, software, methodology, formal analysis, writing original draft, review and editing.

Ying Cai: software, methodology, formal analysis, visualization, writing original draft.

Yun-Jia Wang: software, formal analysis, visualization, writing original draft.

Yue Huang: conceptualization, writing original draft, review and editing.

Chi Zhang: resources, review and editing.

Hu Ma: Supervision, resources, funding acquisition.

Jian-Guo Zhou: conceptualization, methodology, resources, supervision, funding acquisition, project administration, writing original draft, writing–review and editing.

All authors reviewed and approved the final manuscript.

## Declarations

The authors have no competing interests in this work.

## Data Availability

All data produced in the present work are contained in the manuscript
All data produced are available online at http://cancergenome.nih.gov/ and https://www.ncbi.nlm.nih.gov/geo/

