## supplemental figure and tables for "FGF23 is a potential prognostic biomarker in uterine sarcoma"

Supplementary Figures

Supplementary Tables

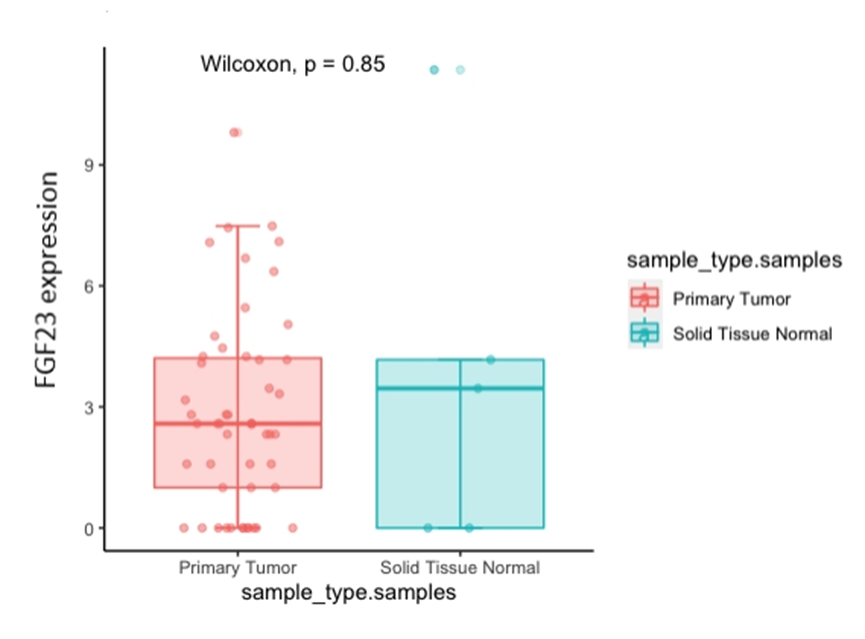

**Supplementary Fig. S1.** The box plot shows the expression of FGF23 between tumor tissue and normal tissue in the TCGA cohort.

**Supplementary Table S1.** Baseline characteristics of 57 UCS patients from the TCGA cohort

| **Characteristics** | **Patients** |
| --- | --- |
| **Age at diagnosis(years)** |  |
| ≤50 | 0 (0%) |
| ＞50 | 57 (100%) |
| **Menopausal status** |  |
| Postmenopausal | 56 (98.25%) |
| Premenopausal | 1 (1.75%) |
| **Race** |  |
| White | 44 (77.19%) |
| Black or African American | 9 (15.79%) |
| Asian | 3 (5.26%) |
| Unknown | 1 (1.75%) |
| **BMI (kg/m^2^)** |  |
| ＜25 | 17 (29.82%) |
| ≥25 | 40 (70.18%) |
| **Peritoneal wash** |  |
| Positive | 12 (21.05%) |
| Negative | 25 (43.86%) |
| Unknown | 20 (35.09%) |
| **FIGO stage** |  |
| Ⅰ | 21 (36.84%) |
| Ⅱ | 6 (10.53%) |
| Ⅲ | 20 (35.09%) |
| Ⅳ | 10 (17.54%) |
| **Surgical approach** |  |
| Minimally invasive | 30 (52.63%) |
| Open | 20 (35.09%) |
| Unknown | 7 (12.28%) |
| **Person neoplasm cancer status** |  |
| With tumor | 9 (15.79%) |
| Tumor free | 11 (19.30%) |
| Unknown | 37 (64.91%) |
| **Adjuvant radiotherapy** |  |
| Yes | 11 (19.30%) |
| No | 9 (15.79%) |
| Unknown | 37 (64.91%) |

**Supplementary Table S2.** Cox univariate and multivariate analysis of the OS of UCS patients from the TCGA cohort

| **Clinicopathological features** | **Univariate analysis** | | | **Multivariate analysis** | | |
| --- | --- | --- | --- | --- | --- | --- |
|  | **HR** | **95% CI** | ***p* value** | **HR** | **95% CI** | ***p* value** |
| **Age (≤65 vs.** ＞65) | 1.04 | 0.98-1.08 | 0.063 | 1.06 | 1.01-1.12 | 0.014 |
| **Tumor stage (I/II vs. III/IV)** | 1.39 | 1.03-1.87 | 0.029 | 1.64 | 1.16-2.31 | 0.005 |
| **Menopausal status (Postmenopausal vs. Premenopausal)** | 2.17 | 0.29-16.20 | 0.449 | 3.95 | 0.39-19.80 | 0.118 |
| **Race (White vs. Other)** | 0.87 | 0.47-1.60 | 0.652 | 0.84 | 0.38-1.81 | 0.651 |
| **BMI (＜25 vs. ≥25)** | 0.71 | 0.36-1.41 | 0.323 | 0.47 | 0.14-1.53 | 0.213 |
| **Surgical approach (Minimally invasive vs. Open)** | 0.81 | 0.37-1.76 | 0.594 | 0.65 | 0.54-2.24 | 0.438 |
| **Person neoplasm cancer status (With tumor vs.** **Tumor free)** | 0.16 | 0.03-0.85 | 0.031 | 0.53 | 0.14-1.32 | 0.004 |
| **Peritoneal wash (Positive vs. Negative)** | 0.19 | 0.07-0.57 | 0.003 | 0.31 | 0.23-0.89 | ＜0.001 |
| **Adjuvant radiotherapy (Yes vs. No)** | 0.46 | 0.26-0.79 | 0.006 | 0.24 | 0.11-0.51 | ＜0.001 |
| **FGF23 expression (High vs. Low)** | 1.06 | 0.47-2.41 | 0.532 | 1.67 | 0.56-2.87 | 0.467 |

**Supplementary Table S3.** Cox univariate and multivariate analyses of the OS in US patients from the Suining cohort

| **Clinicopathological features** | **Univariate analysis** | | | **Multivariate analysis** | | |
| --- | --- | --- | --- | --- | --- | --- |
|  | **HR** | **95% CI** | ***p* value** | **HR** | **95% CI** | ***p* value** |
| **Age (≤50 vs.** ＞50) | 1.03 | 0.99-1.08 | 0.159 | 1.02 | 0.94-1.12 | 0.626 |
| **Tumor stage (I/II vs. III/IV)** | 3.62 | 1.42-9.22 | 0.007 | 0.67 | 0.15-3.11 | 0.611 |
| **Menopausal status (Postmenopausal vs. Premenopausal)** | 1.46 | 0.58-3.69 | 0.424 | 1.94 | 0.40-9.47 | 0.414 |
| **Tumor size (≤5 vs. ＞5)** | 1.85 | 0.66-5.20 | 0.245 | 0.83 | 0.16-4.29 | 0.824 |
| **CA125 (＜35 vs. ＞35)** | 3.23 | 1.58-6.62 | 0.001 | 8.53 | 1.45-50.28 | 0.018 |
| **Tumor type (AS+ESS vs. LMS+UCS)** | 5.71 | 1.30-25.13 | 0.021 | 4.88 | 0.44-54.23 | 0.197 |
| **Lymphadenectomy (Yes vs. No)** | 2.22 | 0.86-5.73 | 0.099 | 2.52 | 0.44-14.42 | 0.300 |
| **Lymphatic metastasis (Yes vs. No)** | 3.09 | 0.40-24.04 | 0.281 | 0.45 | 0.05-3.72 | 0.455 |
| **Tumor recurrence (Yes vs. No)** | 8.47 | 3.08-23.27 | ＜0.001 | 5.59 | 1.30-24.02 | 0.021 |
| **Adjuvant chemotherapy (Yes vs. No)** | 1.14 | 0.45-2.89 | 0.776 | 2.08 | 0.41-10.47 | 0.371 |
| **Adjuvant radiotherapy (Yes vs. No)** | 0.60 | 0.17-2.07 | 0.418 | 0.20 | 0.02-1.64 | 0.134 |
| **FGF23 expression (High vs. Low)** | 1.58 | 0.59-3.99 | 0.383 | 1.37 | 0.79-2.36 | 0.262 |

**Supplementary Table S4.** Cox univariate and multivariate analysis of PFS in US patients from the Suining cohort

| **Clinicopathological features** | **Univariate analysis** | | | **Multivariate analysis** | | | |
| --- | --- | --- | --- | --- | --- | --- | --- |
|  | **HR** | **95% CI** | ***p* value** | | **HR** | **95% CI** | ***p* value** |
| **Age (≤50 vs.** ＞50) | 1.01 | 0.96-1.07 | 0.588 | | 1.04 | 0.94-1.16 | 0.457 |
| **Tumor stage (I/II vs. III/IV)** | 5.51 | 2.02-15.05 | ＜0.001 | | 1.44 | 0.28-7.39 | 0.661 |
| **Menopausal status (Postmenopausal vs. Premenopausal)** | 0.79 | 0.28-2.17 | 0.641 | | 0.33 | 0.04-2.58 | 0.289 |
| **Tumor size (≤5 vs. ＞5)** | 2.97 | 0.84-10.43 | 0.090 | | 3.38 | 0.36-31.97 | 0.288 |
| **CA125 (＜35 vs. ＞35)** | 3.11 | 1.45-6.64 | 0.003 | | 2.29 | 0.77-6.81 | 0.135 |
| **Tumor type (AS+ESS vs. LMS+UCS)** | 10.09 | 1.32-77.08 | 0.026 | | 1.72 | 0.14-21.38 | 0.675 |
| **Lymphadenectomy (Yes vs. No)** | 2.46 | 0.89-6.79 | 0.081 | | 0.99 | 0.18-5.40 | 0.990 |
| **Lymphatic metastasis (Yes vs. No)** | 3.36 | 0.75-15.09 | 0.114 | | 0.39 | 0.05-3.33 | 0.388 |
| **Tumor recurrence (Yes vs. No)** | 20.74 | 5.72-75.14 | ＜0.001 | | 15.29 | 2.60-90.08 | 0.003 |
| **Adjuvant chemotherapy (Yes vs. No)** | 1.54 | 0.57-4.16 | 0.390 | | 3.18 | 0.52-19.38 | 0.209 |
| **Adjuvant radiotherapy (Yes vs. No)** | 0.97 | 0.31-3.03 | 0.968 | | 0.21 | 0.02-1.99 | 0.174 |
| **FGF23 expression (High vs. Low)** | 1.07 | 0.88-1.29 | 0.511 | | 1.13 | 0.79-1.60 | 0.508 |
